# Causal relationship between tea intake and chronic pain: A Mendelian randomization study

**DOI:** 10.1101/2025.05.10.25327365

**Authors:** Shuning Liu, Debin Xu

**Author notes:** Correspondence: Debin Xu, Changchun University of Chinese Medicine, Changchun, Jilin Province, 130117, China.

## Abstract

**Objective:** According to some studies, chronic pain imposes a significant burden on individuals and the economy, affecting more than 30% of the global population. However, the relationship between tea intake and chronic pain remains unclear.

**Methods:** This study employed Mendelian randomization (MR) to detect the causal relationship between tea intake and chronic pain. The tea intake was obtained from the UK Biobank. The Multisite chronic pain (MCP) was used as the primary outcome, while chronic widespread pain (CWP) served as the secondary outcome. To assess heterogeneity, we applied Cochran’s Q statistic with IVW methods. Additionally, the MR-Egger intercept test and MR-PRESSO test were performed to detect potential pleiotropy.

**Results:** The results showed that tea intake increased the risk of MCP. Specifically, an increase in tea intake was associated with a higher risk of MCP (OR = 1.088, 95%CI = 1.038-1.141, P < 0.001). However, no causal relationship was found between tea intake and CWP (OR = 1.006, 95%CI = 0.999-1.014, P > 0.05). Furthermore, no reverse causality was observed.

**Conclusion:** Our findings suggested that genetically predicted tea intake was a risk factor for chronic pain. These results may help shed light on the potential health impacts of tea take, providing further insights into its influence on chronic pain.

## 1. Introduction

According to some studies, chronic pain imposes a significant burden on individuals and the economy, affecting more than 30% of the global population [1]. Unlike acute pain, which serves an essential survival function, chronic pain is increasingly recognized as a distinct disease, with therapeutic (e.g., managing persistent pain) and psychological (e.g., fostering acceptance and optimism) dimensions problem. Chronic pain can be categorized into nociceptive pain (arising from tissue damage), nociplastic pain (resulting from a sensitized nervous system), or neuropathic pain (stemming from nerve damage) [2]. These classifications influence clinical evaluation and treatment decisions. However, in practice, there is considerable overlap in pain mechanisms both within and between patients. This overlap has led many experts to regard pain classification as a continuum rather than discrete categories. Compared to acute pain, chronic pain provides limited, if any, evolutionary advantages [3]. Recognizing chronic pain as a disease allows patients and healthcare providers to shift expectations from eliminating the condition to managing it, focusing on functional recovery and emotional well-being.

Tea, as one of the most widely consumed beverages globally, is rich in bioactive compounds, such as caffeine, tannins, polyphenols, flavonoids, vitamins, saponins, and theanine. These components have been shown to confer various health benefits. For instance, studies attribute the pain-relieving effects of green tea primarily to its antioxidant and anti-inflammatory properties. Phytochemicals in tea can act on immune system cells, antagonize specific cell surface receptors, and suppress the production of inflammatory cytokines, thereby effectively managing pain. Despite these potential benefits, the relationship between tea intake and chronic pain remains debated. Some studies have identified tea intake as a risk factor for syncope in headache patients, while others suggest that tea drinking may elevate the risk of lung cancer and its associated pain. These inconsistent and contradictory findings in traditional epidemiological research highlight the potential influence of confounding factors and reverse causation [4]. Consequently, it remains unclear whether tea intake exacerbates chronic pain or if individuals with chronic pain may turn to tea as a means of alleviation. Clarifying the causal relationship between tea intake and chronic pain is essential. This could inform more effective prevention strategies for managing chronic pain and improving public health outcomes.

Mendelian randomization (MR) is a research approach that leverages genetic variations linked to a specific exposure as instrumental variables (IVs) to detect its causal impact on the risk of particular diseases [5]. We used genetic variations associated with tea intake as IVs to investigate the causal relationships between tea intake and chronic pain both MCP and CWP (Figure 1).

**Figure 1.**
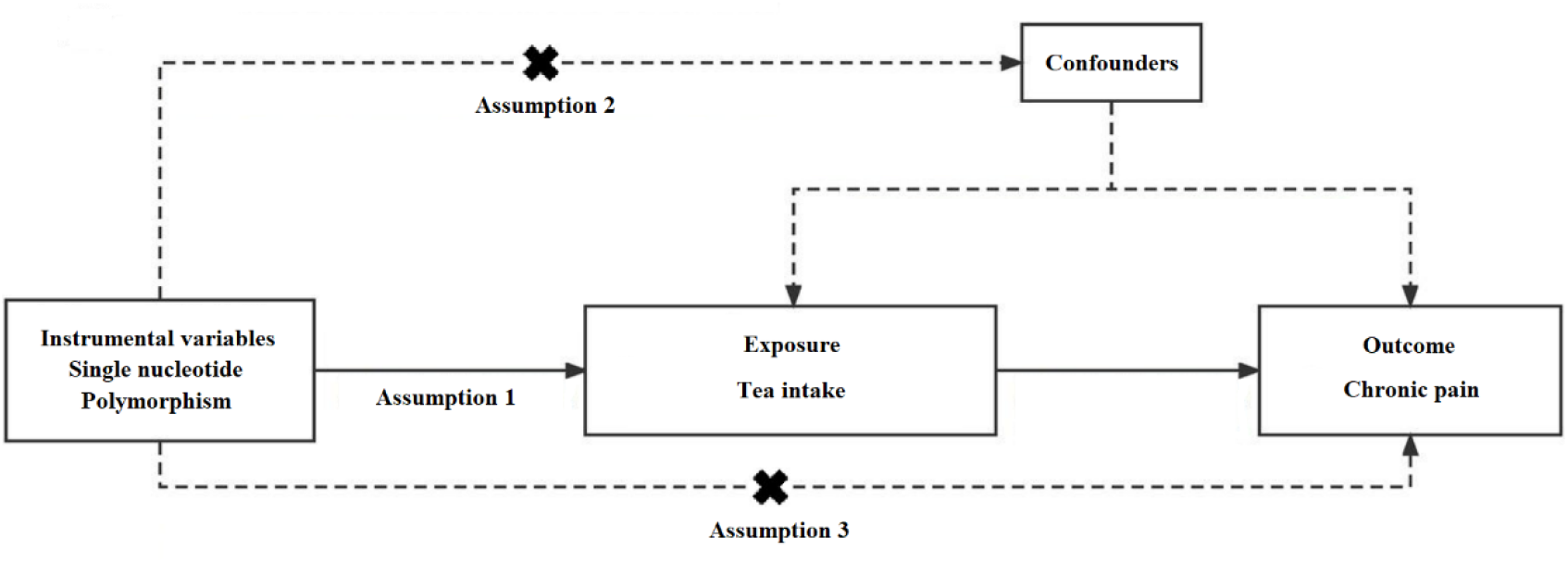
Schematic diagram of MR analysis.

## 2. Materials and Methods

### 2.1. Study design

The MR analysis to explore the causal relationship between tea take and the risk of chronic pain. The primary assumptions of MR are as follows: (a) genetic variation is strongly associated with tea intake; (b) genetic variation is independent of confounding factors; and (c) the effect on chronic pain risk is mediated exclusively through tea intake. Since the analysis utilized publicly available data, it did not require ethical approval or written consent [6].

### 2.2. Data sources

The tea intake was considered the primary exposure, and summary statistics were obtained from the UK Biobank. Data on tea intake were derived from a food frequency questionnaire, with intake quantified continuously as cups per day.

The primary outcome was multisite chronic pain (MCP), with data derived from a GWAS study involving 387,649 European participants, defined as self-reported pain persisting for at least three months in seven specific body regions: the head, face, neck/shoulders, back, stomach/abdomen, hips, and knees [7]. The secondary outcome was chronic widespread pain (CWP), with data from a GWAS studies involving 461,857 European participants. defined as self-reported pain persisting for at least three months in body regions: both above and below the waist, and in the axial skeleton [8] (Table 1).

**Table 1.** Overview of GWAS data.

| Trait | ID | Participants | SNP | Authors | Year |
| --- | --- | --- | --- | --- | --- |
| Tea intake | *ukb-b-6066* | 447,485 | 9,851,867 | Ben E. et al. | 2018 |
| Multisite chronic pain | 33830993 | 387,649 | 9,926,106 | Johnston KJA. et al. | 2021 |
| Chronic widespread pain | 33926923 | 6,914/242,929 | 7,616,783 | Rahman MS. et al. | 2021 |

### 2.3. Instrument selection and data harmonization

We applied a locus-specific significance threshold (P < 5 × 10 ^−6^, r^2^< 0.001, 10,000 kb) [9]. For each IV associated with the exposure trait, we calculated the F-statistic, proportion of explained variance and collected data on key parameters for each SNP. Notably, an F-statistic greater than 10 was deemed indicative of a robust IV [10].

### 2.4. Statistical analyses

We utilized an inverse variance weighting (IVW) method as the primary analytical method, with additional analyses conducted using the Weighted Median, MR-Egger regression, Weighted Mode, and Simple Mode to enhance the robustness of the findings [11]. To assess heterogeneity, we applied Cochran’s Q statistic with IVW methods. The P values greater than 0.05 was considered to indicate no significant evidence of heterogeneity. We executed a “leave-one-out” analysis to evaluate the impact of individual SNP on the causal relationship. Additionally, the MR-Egger intercept test and MR-PRESSO test were performed to detect potential pleiotropy of the IVs, with a P values greater than 0.05 considered evidence of no pleiotropy [12].

## 3. Results

### 3.1. Effect of EA on chronic pain

The results showed that tea intake increased the risk of MCP (Table 2, Figure 2). Specifically, an increase in tea intake was associated with a higher risk of MCP (OR = 1.088, 95%CI = 1.038-1.141, P < 0.001). However, no causal relationship was found between tea intake and CWP (OR = 1.006, 95%CI = 0.999-1.014, P > 0.05). Furthermore, no reverse causality was observed. Specifically, the MCP with tea intake (P = 0.64) and CWP with tea intake (P = 0.27).

**Table 2.** MR results for the effect of tea intake on the chronic pain.

| Exposure | Outcome | Method | OR(CI95%) | $\beta$ | SE | P |
| --- | --- | --- | --- | --- | --- | --- |
| Tea intake | Multisite chronic pain | IVW | 1.088(1.038-1.141) | 0.085 | 0.024 | 4.448e-04 |
|  |  | Weighted Median | 1.14(1.076-1.208) | 0.131 | 0.029 | 8.951e-06 |
|  |  | MR Egger | 1.166(1.027-1.323) | 0.153 | 0.064 | 1.831e-02 |
|  |  | Weighted mode | 1.202(1.112-1.298) | 0.184 | 0.039 | 5.794e-06 |
|  |  | Simple mode | 1.208(1.029-1.417) | 0.189 | 0.081 | 2.141e-02 |
|  | Chronic widespread pain | IVW | 1.006(0.999-1.014) | 0.006 | 0.003 | 6.653e-02 |
|  |  | Weighted Median | 1.006(0.995-1.017) | 0.006 | 0.005 | 2.345e-01 |
|  |  | MR Egger | 1.007(0.989-1.026) | 0.007 | 0.009 | 4.063e-01 |
|  |  | Weighted mode | 1.004(0.978-1.031) | 0.004 | 0.013 | 7.565e-01 |
|  |  | Simple mode | 1.007(0.993-1.02) | 0.007 | 0.006 | 2.856e-01 |

**Figure 2.**
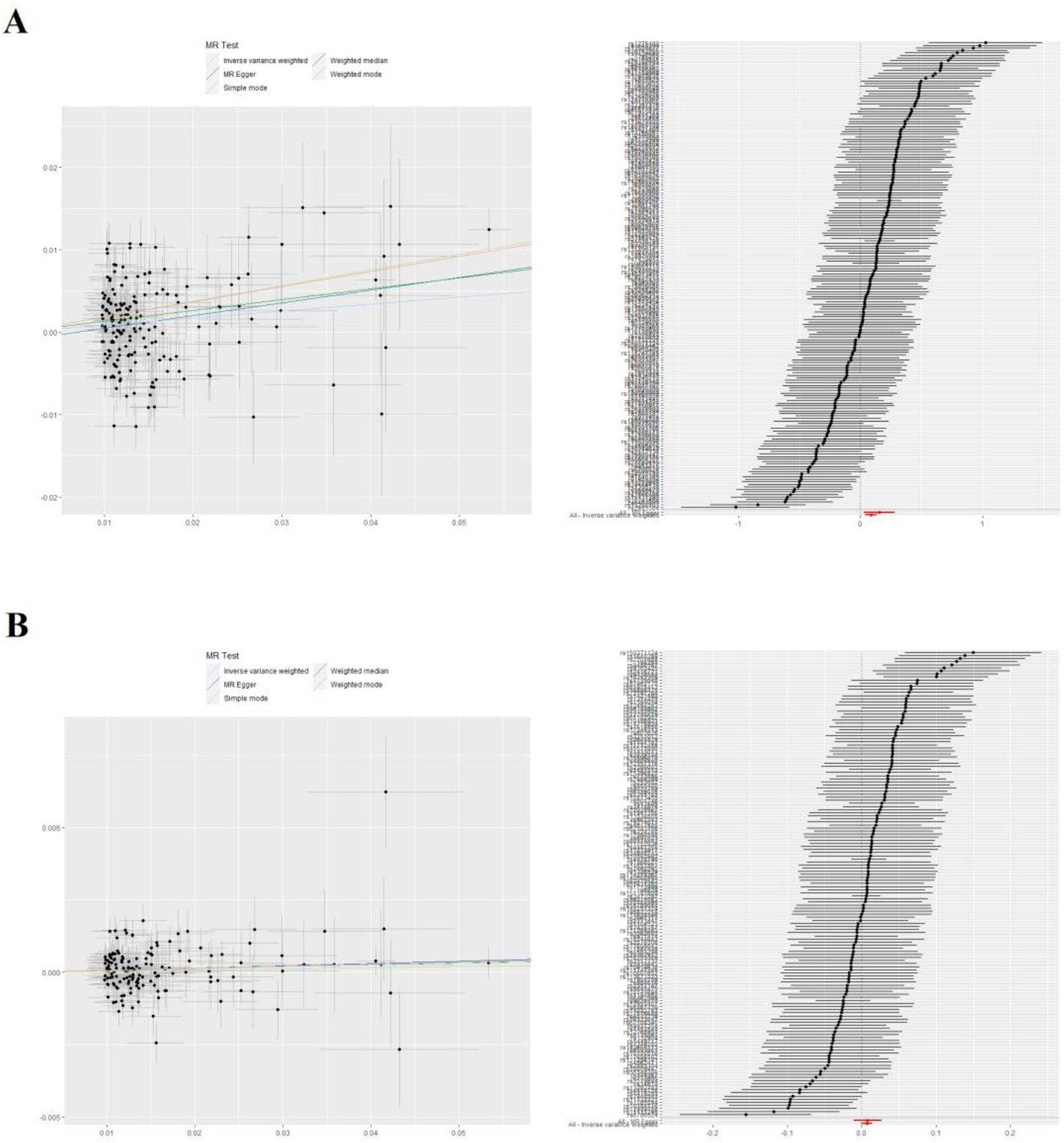
Forest and scatter plot of the MR results, the colored lines depict the fitting results of five MR analysis methods. (A) Multisite chronic pain; (B) Chronic widespread pain.

### 3.2. MR sensitivity analyses

The intercepts of the MR-Egger regression were −0.00165 and 0.00001 (P > 0.01), and the MR-PRESSO analysis showed P values greater than 0.05, suggesting no significant evidence of horizontal pleiotropy in our causal results (Table 3). The Cochrane Q statistics for the IVW method were 488.187 and 215.931 (P < 0.01), which indicated potential heterogeneity in the causal effects of the IVW results; therefore, we applied a random effects model analysis. Furthermore, the funnel plot results displayed general symmetry, providing further evidence that there was little to no heterogeneity or pleiotropy in our causal estimates.

**Table 3.** Sensitivity analysis results.

| Trait | Pleiotropy test |  |  | Heterogeneity test |  |  |
| --- | --- | --- | --- | --- | --- | --- |
| | Intercept | MR-Egger $p$ | PRESSO $p$ | $Q$-statistic | $Q$-$df$ | $Q$-$p$ |
| Multisite chronic pain | -1.065e-03 | 0.252 | 0.258 | 488.187 | 183 | < 0.01 |
| Chronic widespread pain | 1.823e-05 | 0.896 | 0.843 | 215.931 | 150 | < 0.01 |

The “leave-one-out” analysis revealed that excluding any individual SNP did not significantly alter the results, suggesting that the MR analysis findings are not driven by any single SNP. This reinforces the conclusion that the causal relationship between tea take and chronic pain risk is not influenced by individual genetic variants, thereby supporting the robustness of our findings (Figure 3).

**Figure 3.**
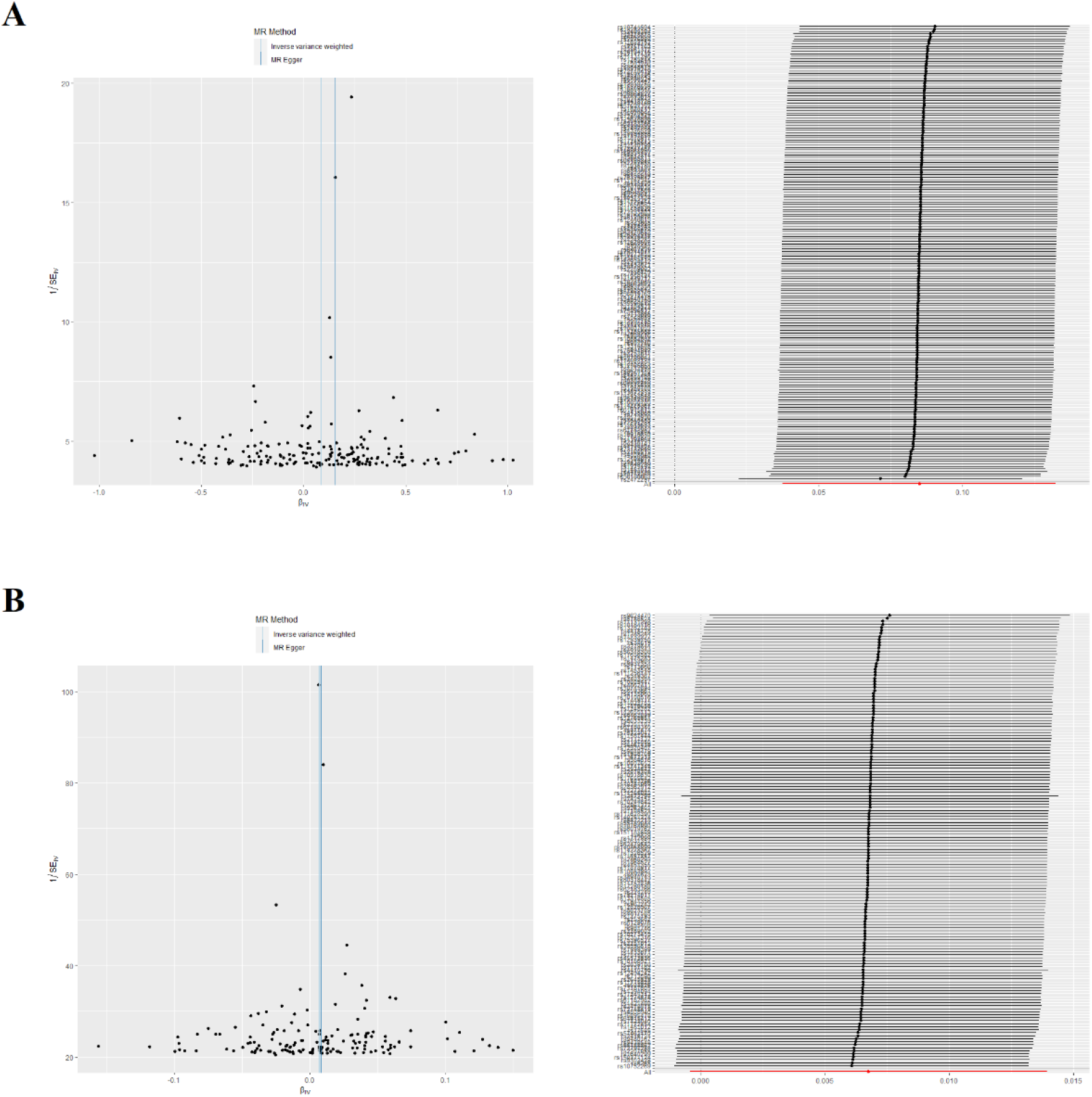
Funnel plot for chronic pain using primary genetic instruments. (A) Multisite chronic pain; (B) Chronic widespread pain.

## 4. Discussion

We identified a significant correlation between higher tea take and an increased susceptibility to chronic pain risk. Notably, this is the first study to utilize the MR approach to systematically investigate the relationship between tea take and chronic pain risk.

Previous studies primarily relied on clinical observations and failed to establish robust causal relationships. For example, one study found that adults consuming green tea beverages exhibited significantly reduced sensitivity to cold pain and a trend toward reduced sensitivity to heat pain [13]. Conversely, another study involving 286 participants reported that tea intake exacerbated their pain symptoms [14]. However, the reliability of these findings is questionable due to limitations such as confounding factors, measurement errors, and reverse causation. One potential source of spurious causal relationships between tea intake and chronic pain is that individuals might use food cravings, including tea, as a coping mechanism to distract themselves from chronic pain episodes [15]. Another explanation is the significant increase in tea intake among chronic pain patients, possibly due to the perception that caffeine serves as an analgesic adjuvant for headache relief, leading to misleading conclusions in traditional studies [16]. Supporting this, our study revealed that higher tea intake might elevate the risk of chronic pain.

Chronic pain is associated with harmful pathophysiological and anatomical changes. These changes may result not only from nociceptive processes but also from psychosocial factors that influence and exacerbate the condition [17]. The biological impacts of chronic pain are diverse and include suppression of both cell-mediated and humoral immunity, changes in gene expression, and reductions in gray matter volume. Many of these detrimental changes can at least partially be reversed through effective treatments, including emotional support systems and interventions that promote physical well-being. Such treatments foster healing and alleviate chronic pain. Similar to other diseases, evidence suggests that appropriate pain management can improve quality-of-life indicators and reverse neuroplasticity changes associated with chronic pain. Given the inherently subjective nature of pain, patient-reported pain experiences should generally be accepted in the absence of conflicting evidence [18]. Physicians may, however, employ additional methods, such as analyzing facial expressions or conducting imaging studies, to assess pain and identify its underlying causes. This underscores the critical importance of pain classification, which influences prognosis, diagnostic evaluation, treatment strategies, healthcare delivery, and prevalence estimates. For instance, in patients with back pain, imaging studies are particularly recommended when invasive surgical interventions are being considered [19]. These are most effective in cases involving neuropathic pain, especially when red flags such as severe or progressive neurological deficits are present, as opposed to non-neuropathic pain.

The continuous advancement of behavioral neuroscience and genome-wide association studies in exploring individualized clinical phenotypes holds significant promise for the development of targeted therapeutic strategies for chronic pain. By enhancing our understanding of the complex interactions between genetics, behavior, and pain mechanisms, these advancements pave the way for more precise and effective interventions [20]. In the perioperative setting, these strategies may include individualized psychological interventions for patients with pre-existing psychopathology, the strategic use of regional anesthesia techniques such as epidural analgesia to prevent persistent pain after high-risk surgeries (e.g., amputations), and preoperative resilience-building measures to enhance postoperative pain management and recovery [21]. The field of epigenetics is also contributing to a deeper understanding of how individual experiences and environmental factors lead to changes in gene expression, altering the function of central nervous system regions implicated in pain chronicity. Current studies are investigating several biomarker categories, including functional and neurochemical imaging, molecular markers (e.g., genomics), psychophysical indicators (e.g., quantitative sensory testing), and behavioral markers (e.g., facial expressions) [22]. Among these, neuroimaging shows particular promise, with advances in specificity achieved through the integration of tools such as multivariate pattern analysis and machine learning.

Although this study offers interesting results into the potential causal relationship between tea intake and the risk of chronic pain, but there are several limitations. First, this study only included European ancestry and was not conducted among other ethnic groups [23]. Second, as tea intake data were self-reported, the possibility of measurement bias cannot be ruled out. Finally, the genetic instruments were selected based on statistical methods rather than biological criteria, potentially reducing their specificity to tea take. This limitation may result in lower heritability estimates for tea take, thereby diminishing the clinical relevance of our MR analysis.

## 5. Conclusions

Our findings suggested that genetically predicted tea intake is a risk factor for chronic pain. These results may help shed light on the potential health impacts of tea take, providing further insights into its influence on chronic pain.

## Data Availability

All data produced in the present work are contained in the manuscript.
UK Biobanks: https://www.ukbiobank.ac.uk
GWAS dataset: https://www.ebi.ac.uk/gwas/studies

## Acknowledgments

The authors thank all participants and investigators who provided the GWAS data.

## Author Contributions

Conceptualization: Shuning Liu; Data curation: Shuning Liu; Methodology: Shuning Liu; Software: Shuning Liu; Visualization: Shuning Liu; Validation: Shuning Liu; Writing—original draft preparation, Shuning Liu; Writing—review and editing: Shunin Liu, Debin Xu; Supervision: Shuning Liu, Debin Xu; Project administration: Debin Xu.

## Funding

This study received no external funding.

## Institutional Review Board Statement

All data are publicly available datasets; therefore, no additional ethical approval was required.

## Conflicts of Interest

The authors declare no conflicts of interest.

## Notes

### Competing Interest Statement

The authors have declared no competing interest.

### Funding Statement

This study did not receive any funding.

### Author Declarations

All data are publicly available datasets; therefore, no additional ethical approval was required.

## References

1. Yong RJ, Mullins PM, Bhattacharyya N. Prevalence of chronic pain among adults in the United States. Pain. 2022; 163: e328–332.

2. Fitzcharles M, Cohen SP, Clauw DJ, et al. Nociplastic pain: towards an understanding of prevalent pain conditions. Lancet. 2021; 397: 2098–110.

3. Clauw DJ, Essex MN, Pitman V, Jones KD. Reframing chronic pain as a disease, not a symptom: rationale and implications for pain management. Postgrad Med. 2019; 131: 185–98.

4. Yong RJ, Mullins PM, Bhattacharyya N. Prevalence of chronic pain among adults in the United States. Pain. 2022; 163: e328–332.

5. Burgess S, Small DS, Thompson SG. A review of instrumental variable estimators for Mendelian randomization. Stat Methods Med Res. 2017; 26(5): 2333–2355.

6. Mou X, Sun M, Chen X. Causal effect of education on bone mineral density: A Mendelian randomization study. Medicine (Baltimore). 2024; 103(11): e37435.

7. Johnston KJA, Adams MJ, Nicholl BI, et al. Genome-wide association study of multisite chronic pain in UK Biobank. PLoS Genet. 2019; 15(6): e1008164.

8. Rahman MS, Winsvold BS, Chavez Chavez SO, et al. Genome-wide association study identifies RNF123 locus as associated with chronic widespread musculoskeletal pain. Ann Rheum Dis. 2021; 80(9): 1227–1235.

9. Hartwig FP, Davies NM, Hemani G, et al. Two-sample Mendelian randomization: avoiding the downsides of a powerful, widely applicable but potentially fallible technique. Int J Epidemiol. 2016; 45: 1717–1726.

10. Shim H, Chasman DI, Smith JD, et al. A multivariate genome-wide association analysis of 10 LDL subfractions, and their response to statin treatment, in 1868 Caucasians. PLoS One. 2015; 10(4): e0120758.

11. Burgess S, Bowden J, Fall T, et al. Sensitivity analyses for robust causal inference from Mendelian randomization analyses with multiple genetic variants. Epidemiology. 2017; 28(1): 30–42.

12. Seblova D, Berggren R, Lövdén M. Education and age-related decline in cognitive performance: Systematic review and meta-analysis of longitudinal cohort studies. Ageing Res Rev. 2020; 58: 101005.

13. Herati AS, Shorter B, Srinivasan AK, et al. Effects of foods and beverages on the symptoms of chronic prostatitis/chronic pelvic pain syndrome. Urology. 2013; 82(6): 1376–1380.

14. Noah L, Morel V, Bertin C, et al. Effect of a Combination of Magnesium, B Vitamins, Rhodiola, and Green Tea (L-Theanine) on Chronically Stressed Healthy Individuals-A Randomized, Placebo-Controlled Study. Nutrients. 2022; 14(9): 1863.

15. Yap ZL, Summers SJ, Grant AR, et al. The role of the social determinants of health in outcomes of surgery for low back pain: a systematic review and narrative synthesis. Spine J. 2022; 22(5): 793–809.

16. Totsch SK, Waite ME, Sorge RE. Dietary influence on pain via the immune system. Prog Mol Biol Transl Sci. 2015; 131: 435–469.

17. Pogatzki-Zahn EM, Segelcke D, Schug SA. Postoperative pain-from mechanisms to treatment. Pain Rep. 2017; 2: e588.

18. Cohen SP, Vase L, Hooten WM. Chronic pain: an update on burden, best practices, and new advances. Lancet. 2021; 397(10289): 2082–2097.

19. Köppen PJ, Dorner TE, Stein KV, et al. Health literacy, pain intensity and pain perception in patients with chronic pain. Wien Klin Wochenschr. 2018; 130(1-2): 23–30.

20. Richebé P, Capdevila X, Rivat C. Persistent postsurgical pain: pathophysiology and preventative pharmacologic considerations. Anesthesiology. 2018; 129: 590–607.

21. Cohen SP, Wallace M, Rauck RL, et al. Unique aspects of clinical trials of invasive therapies for chronic pain. Pain Rep. 2018; 4: e687.

22. Dale R, Stacey B. Multimodal Treatment of Chronic Pain. Med Clin North Am. 2016; 100(1): 55–64.

23. Ma X, Sun J, Geng R, et al. Depression and the risk of fibromyalgia syndrome: a two-sample Mendelian randomization study. Front Psychiatry. 2024; 15: 1282172.

